# Higher SARS-CoV-2 Transmission Burden Among Racialized Individuals: Evidence from Canadian Serology Data

**DOI:** 10.64898/2026.03.23.26349092

**Authors:** Simran Mann, Natalie Wilson, Clara Lee, David N. Fisman

**Author notes:** **Address reprint requests and correspondence to:** David Fisman, MD MPH FRCP(C), Room 686, 155 College Street, Toronto, Ontario, Canada, M5T 3M7. **Data Availability Statement** **Code Availability:** Analysis code is available at https://github.com/fismanda/covid19-seroprev-race and permanently archived at https://doi.org/10.5281/zenodo.18818120. **Data Availability:** Seroprevalence data used in this study are publicly available from the COVID-19 Immunity Task Force Data Portal at https://portal.citf.mcgill.ca.

## Abstract

**Introduction:** COVID-19 transmission has not been evenly distributed across racial groups, with exposure being shaped by social and structural factors. The emergence of highly transmissible variants (i.e., Omicron) dramatically increased infection rates. However, it remains unclear whether racial disparities in transmission disappeared or persisted over the course of the pandemic.

**Objective:** To understand how SARS-CoV-2 transmission differed by race in Canada and whether those disparities changed with the Omicron variant.

**Methods:** We analyzed cross-sectional SARS-CoV-2 seroprevalence data from the Canadian Blood Services serosurveillance program (June 2020 to April 2023) using a previously described dynamic susceptible-infection model, while accounting for seroreversion. Race-specific force of infection was estimated for the pre-Omicron and Omicron periods (with the emergence of Omicron defined as beginning December 26, 2021).

**Results:** Prior to Omicron, racialized individuals had a 74% higher force of infection (IRR = 2.205; 95% CI: 2.115-2.299). During the Omicron period, infection rates rose significantly within each racial group relative to the pre-Omicron period, with a 55.52-fold increase among White individuals and a 31.27-fold increase among racialized individuals. Despite this, racialized individuals remained disproportionately affected following the emergence of Omicron, with 24% higher infection rates than those of their White counterparts (IRR = 1.242; 95% CI: 1.231-1.253).

**Conclusion:** Widespread transmission during Omicron did not result in epidemiologic equity, as racialized populations continued to experience higher infection risk despite crude seroprevalence depicting convergence.

## Introduction

The severe acute respiratory syndrome coronavirus 2 (SARS-CoV-2) pandemic highlighted many social and structural health inequities globally. Across high-income countries, racialized populations were disproportionately affected, being subjected to higher rates of COVID-19 infections, hospitalizations and mortality (1, 2). In Canada, despite access to a universal healthcare system, racialized individuals faced elevated risks of SARS-CoV-2 infections and more adverse health outcomes, particularly among Black and South Asian populations (3–5). Early public health surveillance further demonstrated higher COVID-19 case rates, hospitalizations, and deaths in ethnically diverse neighbourhoods compared to less diverse areas (6, 7). Together, this evidence underscores the persistence of racial inequities in COVID-19 outcomes and the need to examine transmission dynamics through an equity lens.

Race, a social construct, is operationalized to categorize individuals by physical differences (3, 8). In this context, the term race is used to describe racialized populations and does not encompass Indigenous persons, whose health inequities are distinct and rooted in colonialism (3). Racialized populations experience unique structural barriers and systemic racism that shape income security, access to paid sick leave, and overrepresentation in front-line work (3). These factors limit their ability to engage in public health interventions (e.g., social isolation, remote work), resulting in greater susceptibility to COVID-19 infections and poorer health outcomes (9).

The emergence of the Omicron variant in December 2021 marked a distinct epidemiologic period, as it altered both transmission and immune protection. Compared to preceding variants, Omicron’s increased transmissibility and immune evasion resulted in a rapid surge in global infection rates (10), and in Canada, this shift coincided with a notably larger increase in cases among those vaccinated compared to the pre-Omicron period (11). At the same time, Canadian race-based COVID-19 data indicate lower vaccine uptake among Arab, Latin American, Black, and Indigenous communities compared to non-visible minorities (12). Prior work suggests these differences stem from structural factors, including mistrust in health care institutions due to colonialism, experiences of discrimination, misinformation, literacy, and systemic racism (13). Complementary evidence from the United States further illustrates these differences (1). Although hospitalization rates increased across all groups during the Omicron period when compared to the Delta-predominant period, irrespective of vaccination status, unvaccinated Black adults experienced a disproportionate burden of hospitalizations (1). Together, these patterns suggest that differential vaccine uptake across racial groups may have influenced infection dynamics during the Omicron period.

Official case counts underestimate infections as they rely on health-seeking behaviours and overlook asymptomatic individuals, making it difficult to assess true racial disparities (14, 15). Serosurveillance addresses this limitation by measuring antibodies from prior infections, providing a more complete estimate of cumulative infection (14, 16). In Canada, the COVID-19 Immunity Task Force (CITF) offers seroprevalence data using nucleocapsid protein detection to identify prior infections (17). Yet seroprevalence may obscure differences in transmission due to antibodies waning (seroreversion) over time, and few studies model this effect (14,16). Therefore, estimating the force of infection (FOI), a dynamic measure of the rate at which susceptible individuals become infected (or, when an antibody is not protective against infection, the rate at which seronegative individuals become seropositive), enables a clearer comparison of transmission dynamics (18).

Recent work by Hassan et al. (15) used CITF data to examine the effects of neighbourhood-level material deprivation on COVID-19 force of infection. They found that apparent convergence in seroprevalence across socioeconomic strata during the Omicron period masked persistent disparities in FOI. Their findings suggested that crude seroprevalence convergence may be a mathematical artifact of differential infection saturation rather than evidence of reduced inequity. Building on this work, this study applies Hassan et al’s (15) modelling framework to race-stratified seroprevalence CITF data. The objectives of this study are to: (1) quantify racial differences in SARS-CoV-2 force of infection in Canada, and (2) assess whether transmission patterns across racial strata changed after the emergence of the Omicron SARS-CoV-2 variant in December 2021.

## Methods

### Data Sources and Study Population

We conducted a cross-sectional study using aggregated population-level SARS-CoV-2 seroprevalence data from the CITF. For the purposes of this study, analysis was restricted to include data from the Canadian Blood Services (CBS) serosurveillance program and seroprevalence estimates of antibodies targeting only the nucleocapsid protein (anti-N). These antibodies indicate prior infections but are not elicited in response to COVID-19 vaccination, which targets the spike protein (17, 19–21). Thus, anti-N antibodies provide a reliable marker for past SARS-CoV-2 infections.

The study population included adult blood donors (≥17 years) who met the CBS blood donation eligibility criteria. Individuals from Quebec and the Northern territories (Yukon, Northwest Territories, and Nunavut) are not included in the analysis, as CBS does not collect blood donations in these provinces (19). Additionally, blood donors represent a healthier portion of the population, otherwise known as the healthy donor effect, thus impacting the representativeness of these findings (22).

### Study Period and Definitions

The model was initialized to March 2020, coinciding with the onset of community transmission in Canada (23). Analyses were stratified into two time periods reflecting major pandemic phases: (1) Pre-Omicron, including the Alpha, Beta, Gamma and Delta variants from June 14, 2020, to December 25, 2021; and (2) Omicron, from December 26, 2021, to April 26, 2023 (24).

Racial identity information was self-reported in the CBS dataset and was dichotomized into ‘White’ and ‘racialized’ categories by Swail et al. (25) for analytic purposes. The binary nature of race-based data limits granularity and may mask differences among diverse racialized populations, thereby reducing generalizability. This analysis included 193 total seroprevalence observations (96 White, 97 racialized), after exclusion of one erroneous observation (see below). Each observation corresponds to one cross-sectional CBS seroprevalence estimate collected between June 2020 and April 2023. Sample denominators per observation ranged from 996 to 52,852 blood donors.

### Statistical Analysis

Seroprevalence data reflects cumulative infections; however, antibody waning may lead to seroreversion. To account for this, seroconversion and seroreversion rates were estimated using the same mathematical model previously described by Hassan et al (15).

The model is defined by:

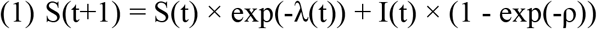

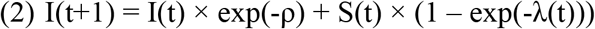

where S(t) = proportion susceptible (seronegative) at time t, I(t) = proportion infected (seropositive) at time t, λ(t) = time-varying force of infection, and ρ = seroreversion rate (per capita rate of antibody loss).

This study adopted an identical model structure, including the saturated and parsimonious model specifications as Hassan et al. (15), to estimate SARS-CoV-2 FOI over time by racial groups. In the parsimonious model, baseline forces of infection were specified by race with a shared Omicron multiplier, whereas the saturated model allowed Omicron’s transmission to differ across racial groups, estimating separate forces of infection for each time period. Key assumptions at model initialization were that all individuals were susceptible (S(0) = 1) and no active infections were present (I(0) = 0). Parameter estimation began in June 2020, when race data became available, using maximum likelihood estimation. We evaluated model fit using the same criteria (Akaike information criterion, Bayesian information criteria, and likelihood ratio tests). Model selection between the saturated and parsimonious models was conducted through the same procedures and the saturated model was retained based on superior fit.

All analyses were conducted using R version 4.4.3. Weekly and absolute FOI estimates were calculated for each racial group during both pre-Omicron and Omicron periods. Incidence rate ratios (IRRs) with 95% confidence intervals (CIs) were computed to compare FOI between White and racialized groups. To assess whether the emergence of the Omicron variant led to differential amplification of transmission by race, an Omicron multiplier was calculated for each racial group as the ratio of FOI before and after the Omicron period. Key R packages used included bbmle, dplyr, and ggplot2.

### Ethical Considerations

This study was approved by the University of Toronto Health Sciences Research Ethics Board (Protocol #46160). The requirement for informed consent was waived because the study used de-identified, aggregated seroprevalence data publicly released by the COVID-19 Immunity Task Force. No individual-level data were accessed. Ethical approval for the original serosurveillance program was obtained by CBS and participating institutions, as previously described (19, 20).

## Results

The analysis includes 193 total observations (96 White, 97 racialized) spanning from June 2020 to April 2023, after exclusion of one erroneous observation in the White group (December 28, 2021; reported seroprevalence 89.6%) due to apparent transposition of anti-N and anti-S values (surrounding anti-N values were approximately 5%). Seroprevalence denominators ranged from 996 to 52,852. The saturated model allowing race-specific Omicron effects demonstrated an improved fit, compared to the parsimonious model, as indicated by a lower AIC (ΔAIC = −551.2) and a significant likelihood ratio test (p = 2.53 x 10^-122^). The saturated model estimated a seroreversion rate of 0.116% per week and a half-life of 596.2 weeks (137.2 months), indicating minimal antibody waning. A temporary visual convergence in seroprevalence was observed at the onset of the Omicron period (late 2021 to early 2022), after which disparities between racial groups re-emerged (**Figure 1**).

**Figure 1.**
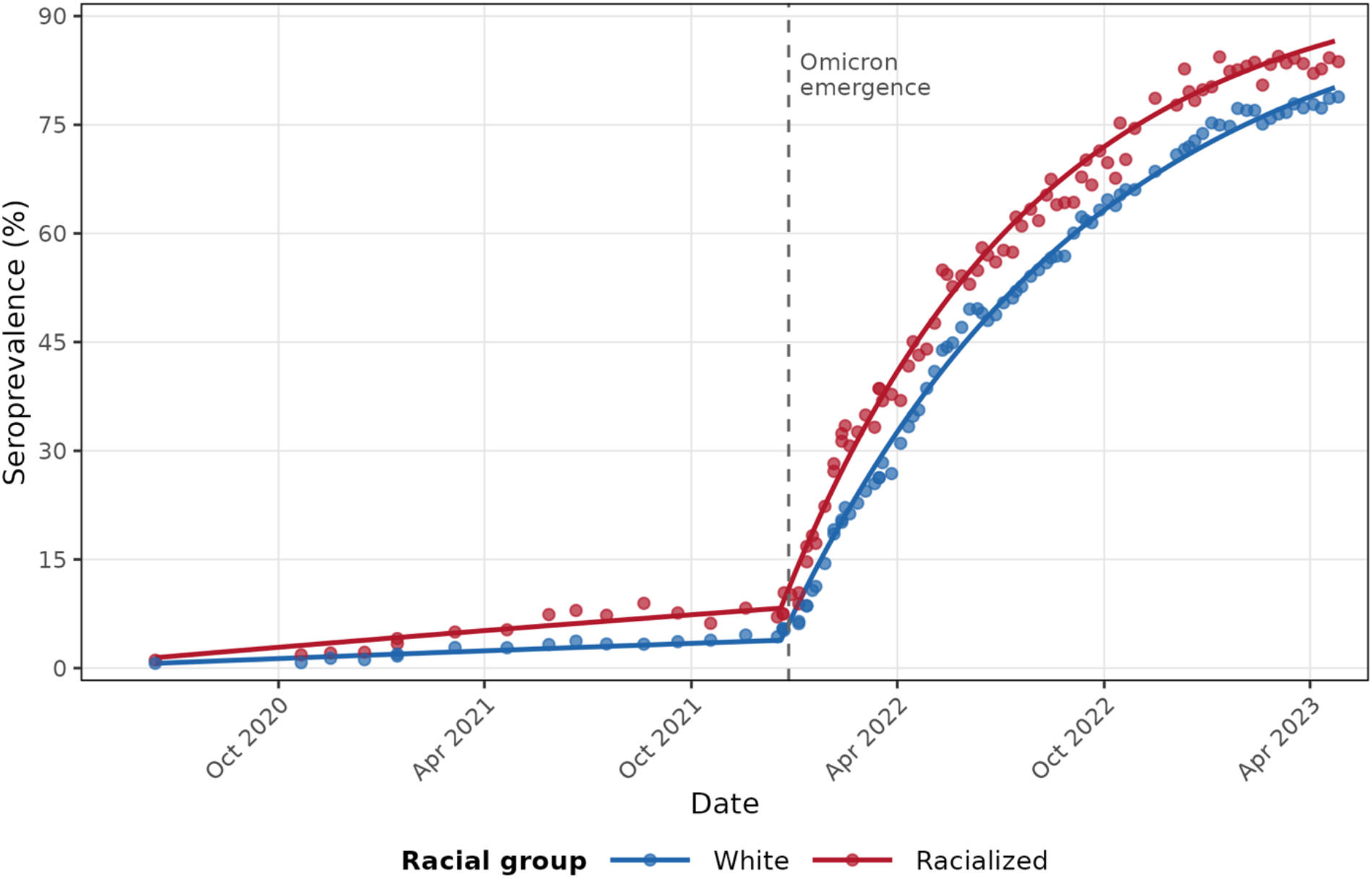
Infection-induced SARS-CoV-2 seroprevalence by racial groups (White and racialized) in Canada, June 2020 to April 2023. Observed (points) and model-predicted (lines) seroprevalence are shown. Seroprevalence increases over time and rises rapidly following the emergence of Omicron (dotted vertical line). A temporary visual convergence of seroprevalence estimates for racial groups is observed in late 2021 and early 2022.

Estimated weekly FOI differed by racial group in both time periods (Table 1). In the pre-Omicron period, weekly FOI among White individuals was 0.00044 and 0.00097 among racialized individuals, corresponding to a higher FOI among racialized (IRR=2.205; 95% CI: 2.115-2.299). During the Omicron period, weekly FOI increased substantially in both groups, reaching 0.02442 among White individuals and 0.03032 among racialized individuals. Although racialized individuals continued to experience a higher FOI, relative difference between groups was smaller than in the pre-Omicron period (IRR = 1.242, 95% CI: 1.231-1.253).

**Table 1:** Saturated Model Pre-Omicron and Omicron Force of Infections (per week) and Incidence Rate Ratios, with 95% Confidence Intervals, by Racial Groups in Canada.

|  | **Racial Group** | **Weekly FOI** | **IRR (95% CI)** |
| --- | --- | --- | --- |
| **Pre-Omicron** | White | 0.00044 | 1.000 (ref) |
|  | Racialized | 0.00097 | 2.205 (2.115, 2.299) |
| **Omicron** | White | 0.02442 | 1.000 (ref) |
|  | Racialized | 0.03032 | 1.242 (1.231, 1.253) |

Abbreviations: FOI, Force of Infection; IRR, Incidence Rate Ratio; CI, Confidence Interval; ref, referent.

The transition to the Omicron period was associated with a larger increase in weekly FOI across both racial groups (Table 2). Among White individuals, the absolute increase in weekly FOI was 0.02398, corresponding to a 55.52-fold increase relative to the pre-Omicron period (95% CI: 54.05-57.03). Among racialized individuals, the absolute increase in weekly FOI was 0.02935, corresponding to a 31.27-fold rise (95% CI: 30.09-32.49). Differential amplification led to the compression of the racial disparity in FOI, as reflected by the reduction in IRRs for racialized individuals (2.205 pre-Omicron to 1.242 during Omicron) and an Omicron multiplier ratio (White/racialized) of 1.78. Despite this compression, racialized individuals continued to experience higher FOI during the Omicron period.

**Table 2:** Race-Specific Force of Infection (per week) and Omicron Multiplier with 95% Confidence Intervals for the Saturated Model.

| **Racial Group** | **Absolute Weekly FOI** | **Omicron Multiplier** | **95% CI** | **Ratio\*** |
| --- | --- | --- | --- | --- |
| White | 0.02328 | 55.52x | 54.05, 57.03 | 1.78 |
| Racialized | 0.02892 | 31.27x | 30.09, 32.49 |  |

Abbreviations: FOI, Force of Infection; CI, Confidence Interval.
\*Ratio = White/Racialized

## Discussion

This study found that there were differences in FOI across racial groups, although the emergence of Omicron compressed these disparities. Before Omicron, the FOI for the racialized groups was significantly higher (IRR=2.205; 95% CI: 2.115-2.299), but during Omicron the IRR decreased to 1.242 (95% CI: 1.231-1.253) when compared to their White counterparts, representing a reduction of 44%. However, this compression should not be interpreted as epidemiologic equity in transmission. Dynamic FOI modeling demonstrated that the narrowing IRR resulted from a larger proportional increase in FOI among White individuals, while racialized groups continued to experience higher absolute FOI. Thus, Omicron intensified transmission broadly but did not reduce structural inequities in exposure.

Hassan et al. (15) argued that apparent seroprevalence convergence in COVID-19 across socioeconomic strata during the Omicron period was not evidence of reduced transmission inequities, but rather a mathematical consequence of differential amplification and a decreasing seronegative denominator of individuals “at risk” of infection, which meant that similar absolute fractions of the population infected in disadvantaged and advantaged individuals would imply far higher FOI among the (more commonly anti-N seropositive) disadvantaged population. The findings of this analysis align closely with this framework and extends it to racial stratification.

In crude seroprevalence data, both White and racialized individuals reach high cumulative seroprevalence by the end of the study period (approximately 70-85%), which could be interpreted as convergence. However, the dynamic FOI estimates reveal persistent disparities where racialized individuals continued to experience higher FOI in both pre-Omicron and Omicron periods. The narrowing of relative differences during Omicron (1.73 to 1.25) was driven by a larger proportional increase in FOI among White individuals rather than a reduced transmission among racialized groups. This supports Hassan et al’s (15) conclusion that seroprevalence is likely a saturation-driven artifact. As the racialized populations have higher infection burden early on during the pandemic, they had fewer susceptible individuals remaining during the emergence of Omicron, limiting further increases in seroprevalence despite elevated transmission.

Consistent with prior analyses of the CBS serosurveillance data, we observed higher infection transmission among racialized individuals prior to the emergence of Omicron. In the Omicron period, racialized populations continued to experience higher infection rates; however, with smaller effect sizes (26). While earlier studies, which focused primarily on temporal trends, attributed attenuated sociodemographic differences to Omicron’s heightened transmissibility (26), our race-centered analyses demonstrates that the apparent convergence reflects a rapid escalation of infection among White populations rather than a reduction in infection burden among racialized groups. Differential vaccination coverage may have contributed to persistent FOI disparities in racialized individuals later in the pandemic period, but it does not explain the compression observed during Omicron, which was driven primarily by a rapid FOI increase among White individuals with larger remaining susceptible pools. However, the persistence of higher FOI among racialized groups is consistent with continued inequities in exposure and protection, suggesting that structural factors remain integral contributors to disparities even as overall transmission increased.

### Limitations

Due to reliance on blood donor serology data and the CBS eligibility criteria, those who donate blood exhibit a “healthy donor effect”, meaning they tend to be healthier and have healthier lifestyles than the general population (22). Second, race was dichotomized into White versus racialized categories, reducing granularity and preventing examination of specific trends within different racial groups. Additionally, this classification does not fully capture Indigenous identity (3), and it is unclear how Indigenous individuals are represented within the CITF data’s racialized category. Lastly, race is intertwined with multiple structural determinants (e.g., immigration status, housing density, occupation, access to healthcare), and observed disparities likely reflect intersecting systems of disadvantage rather than race alone. Due to the aggregated nature of the data, these factors could not be directly accounted for in this analysis, limiting causal interpretation and highlighting the need for intersectional approaches in future research.

### Future Research

Future work should incorporate more detailed race categories and intersectional lens to capture individual-level socioeconomic and structural variables to better explain underlying drivers of transmission disparities. Further studies should apply this dynamic compartmental model to estimate disparities in COVID-19 related health outcomes, such as severity of infection, hospitalizations, and reinfection risk, to provide a more comprehensive understanding of racial inequities across the pandemic.

## Conclusion

Our findings demonstrate that racialized individuals in Canada experienced higher SARS-CoV-2 force of infection than White individuals throughout the pandemic. Although convergence was observed during the beginning of Omicron period, disparities in transmission continued to persist. This work emphasizes the value of dynamic modelling for seroprevalence data, offering a more nuanced interpretation than static comparisons and highlights how convergence in infection rates does not necessarily equate epidemiologic equity. These results can inform future outbreak preparedness by supporting equity-focused resource allocation and interventions that reach high-risk populations effectively.

## Data Availability

Code Availability: Analysis code is available at https://github.com/fismanda/covid19-seroprev-race and permanently archived at https://doi.org/10.5281/zenodo.18818120. Data Availability: Seroprevalence data used in this study are publicly available from the COVID-19 Immunity Task Force (CITF) Data Portal at https://portal.citf.mcgill.ca.

## Declarations

### Ethics approval and consent to participate

This study was approved by the University of Toronto Health Sciences Research Ethics Board (Protocol #46160). The requirement for informed consent was waived by the Research Ethics Board because the study used pre-collected/secondary data.

### Consent for publication

Not applicable.

### Authors’ contributions

SM and DNF designed the study. DNF conceived the work and acquired the data. SM, NJW, CEL, and DNF contributed to analysis and interpretation of data and drafted and/or substantively revised the manuscript. All authors read and approved the final manuscript and take responsibility for the submitted work.

## Acknowledgements

Not applicable.

## Authors’ information (optional)

DNF is the corresponding author: David Fisman MD MPH

